# Assessing Diagnostic Performance of Plasma Biomarkers in Alzheimer’s Disease: P-tau217 Emerges as the Optimal Marker in Chinese Cohorts

**DOI:** 10.1101/2024.12.25.24319645

**Authors:** Shinan Wang, Dequan Liu, Haiyan Li, Xiaodong Jia, Hailan Zhou, Wenying Yu, Tong Li, Liping Pan, Baorong Chen, Yujia Wang, Nan Zhan, Yijun Song, Keqiang Yan

## Abstract

The Simoa platform is recognized as a highly sensitive tool for detecting blood-based biomarkers of Alzheimer’s disease (AD). It is extensively utilized in the diagnosis and identification of AD, with accuracy emerging as a pivotal metric for assessing assay performance, gradually gaining acceptance and application. The primary objective of this study was to assess the diagnostic efficacy of multiple biomarkers in AD using the Simoa platform. The ultimate goal was to identify the optimal diagnostic biomarkers and further investigate their practical application value in the Chinese population. The study comprised two cohorts: cohort I consisted of 151 healthy controls and 90 AD patients, while cohort II was sourced from a Chinese population cohort, encompassing 123 healthy controls and 126 AD patients, utilizing publicly available data. All patients underwent plasma biomarker concentration measurements using the Simoa platform. The specificity, sensitivity, and accuracy of these biomarkers for AD diagnosis were compared to evaluate their diagnostic efficacy. The findings revealed that plasma P-tau217 exhibited excellent performance in differentiating AD from healthy controls, with a sensitivity of 95.0%, specificity of 96.0%, and accuracy of 95.7% for AD diagnosis. Conversely, other indicators, including Aβ42, Aβ42/40, T-tau/Aβ42, and P-tau181, demonstrated some diagnostic efficacy but fell short of meeting the diagnostic criteria. P-tau217 stands out as a highly effective biomarker for the diagnosis of AD, exhibiting extensive clinical application potential in the Chinese population. It presents a promising array of clinical prospects for the Chinese population.

## Introduction

Alzheimer’s disease (AD) represents the primary cause of dementia, imposing a substantial social and economic burden on global societies^[1]^. The acceleration of global population aging has led to a surge in AD prevalence, anticipating to affect over 100 million individuals by 2050. This phenomenon has emerged as a significant global challenge and poses a grave threat to societal structures and healthcare systems^[2]^. As China has officially transitioned into a phase of profound population aging, the incidence of AD has markedly increased, rendering related issues increasingly prominent. The China Alzheimer’s Disease Report 2024 indicates that the prevalence and mortality rates of AD and other forms of dementia in China escalate significantly with age, with significant gender differences^[3]^. Specifically, the prevalence rate among women stands at 1558.9 per 100,000, with a mortality rate of 47.4 per 100,000, whereas for men, the corresponding rates are 846.3 per 100,000 and 22.5 per 100,000, respectively. These statistics underscore the heightened risk of AD in women and underscore the critical need for targeting this demographic for early screening and intervention strategies. Cognitive impairment, a clinical condition that often receives inadequate attention, is predominantly observed within communities or at the grassroots level. However, only a fraction of these cases are referred to memory clinics or specialized hospitals for comprehensive evaluations and consultations^[4]^.

Recent findings indicate that early intervention in at-risk populations is efficacious in mitigating the risk of developing AD^[5]^. Additionally, the emergence of new therapies emphasises the need for sensitive and reliable tests to identify patients who may benefit from early intervention^[6, 7]^. Traditional face-to-face neuropsychological assessments typically detect changes only after several years of amyloid and tau pathology accumulation, which may have contributed to the failure of previous clinical trials^[8]^. Within the diagnostic paradigm for AD in the laboratory setting, cerebrospinal fluid (CSF) analysis and Aβ/Tau-PET imaging are regarded as the gold standard^[9]^. Amyloid β (Aβ) and tau proteins in cerebrospinal fluid, quantified through positron emission tomography (PET) imaging, serve as indicators of the core pathology of AD, and these biomarkers are universally acknowledged. However, these diagnostic tools are frequently invasive, costly, and in certain instances, difficult to procure, leading to a misdiagnosis of symptomatic Alzheimer’s disease in 25% to 35% of patients managed in some cognitive impairment clinics^[10, 11]^. Consequently, the scarcity of user-friendly biomarker assays for Alzheimer’s disease poses a significant impediment to the initiation and effective deployment of anti-amyloid immunotherapy for the treatment of patients with Alzheimer’s disease^[12]^.

In recent years, an escalating body of evidence has indicated that blood-based biomarkers exhibit considerable promise for screening, early diagnosis, monitoring disease progression, and evaluating treatment efficacy in AD^[13-16]^. Notably, the implementation of ultrasensitive assays, such as the Simoa platform, has emerged as a viable alternative to cerebrospinal fluid testing for diagnostic purposes^[17]^. Among the various available platforms, the Simoa platform stands out as the clinically preferred option due to its exceptional stability and performance. According to the updated guidelines from the International Conference on Alzheimer’s Disease in 2023, all biomarkers must meet stringent accuracy requirements when assessing their clinical diagnostic utility^[18]^. However, comparative studies examining the diagnostic efficacy of multiple biomarkers in AD, particularly those based on the Simoa platform, remain scant in the Chinese population. This has led to a lack of a solid basis for clinical selection, resulting in increased patient burden due to repeated testing. Therefore, there is an urgent need for in-depth studies to address this gap. Our study aims to provide ideal biomarker options for clinical application, thereby alleviating the economic burden on patients and facilitating early and accurate diagnosis of AD.

## Methods

### Study design and participants

This study conducted a comprehensive analysis of data from two cohorts. Cohort I consisted of patients recruited from the Memory Clinic at Tianjin Medical University General Hospital, encompassing 90 cases of AD. These patients, aged between 42 and 85, were diagnosed by professional physicians using specific IWG-2criteria. Additionally, this study enrolled 151 cognitively unimpaired (CUC) healthy individuals from a clinical laboratory center in the Northern China, Hainan General Hospital in the Southeast China, and the Affiliated Hospital of Guizhou Medical University in the Southwest China. These participants exhibited no subjective symptoms of cognitive decline, demonstrated normal neurological and neuropsychological examination results, had MMSE scores greater than 26, CDR scores of 0, and ranged in age from 21 to 85 years. Exclusion criteria for both the patients and CUC healthy individuals encompassed active substance abuse, alcoholism, recent head trauma, major recent surgeries, tumors, multiple sclerosis, hydrocephalus, schizophrenia, thyroid dysfunction, vitamin B12 deficiency, abnormal renal function, syphilis or HIV infection, severe depression, or visual/hearing impairments. Moreover, patients receiving disease-modifying treatments or involved in clinical trials were excluded. The study received approval from the Medical Research Ethics Committee of the General Hospital of Tianjin Medical University (Ethics Number: IRB2023-YX-214-01). All participants provided written informed consent at recruitment. The data pertaining to cohort II originated from a study on AD within the Chinese population, undertaken by Xuanwu Hospital^[19]^. Utilizing the Simoa platform, the research aimed to detect levels of Aβ40, Aβ42, Tau, and P-tau181 in the plasma of 123 healthy controls and 126 individuals diagnosed with AD.

### Plasma Aβ40, Aβ42, P-tau217 and Tau measurement

Sample collection involved using an EDTA anticoagulant tube to obtain 5ml of peripheral venous blood from the subjects. The collected blood was centrifuged at 3000r/min for 5 minutes in a high-speed centrifuge, and the plasma was separated and stored at -80°C for further analysis. Plasma Aβ42, Aβ40, Aβ42/40, T-tau, and P-tau217 levels were measured using the fully automated Simoa HD-1 protein detection analyzer (Quanterix Corporation, USA), which utilizes Simoa technology. This technology is a bead-based experimental approach that is 1000 times more sensitive than the traditional enzyme-linked immunosorbent assay (ELISA). All procedures were conducted in strict accordance with the manufacturer’s instructions.

### Statistical analysis

Normally distributed measurement data were presented as mean ± standard deviation and subjected to t-tests, while non-normally distributed measurement data were described using median and interquartile range and assessed through nonparametric tests: Kruskal-Wallis or Wilcoxon tests. Sensitivity and specificity analysis of discrimination was expressed by receiver operating characteristic (ROC) curve and area under the curve (AUC). Boxplots and points were used to present the distributions of original values of plasma biomarkers. All data were analyzed using R software.

## Results

### A Differential Analysis of Each Indicator Across Various Groups

According to the results of Cohort I, significant disparities were observed in the concentrations of P-tau217, Aβ42, Aβ40, Aβ42/Aβ40 ratio, Tau, and Tau/Aβ42 ratio between the AD group and the control group (P < 0.05, Table 1). These findings align with existing research on the use of biomarkers in disease diagnosis. Specifically, the AD group exhibited notably higher concentrations of P-tau217, Aβ40, Tau, and Tau/Aβ42 ratio compared to the control group. In contrast, the control group had higher concentrations of Aβ42 and the Aβ42/Aβ40 ratio (Figure 1). Upon examining the biomarkers in Cohort II, similar significant differences were found in the concentrations of P-tau181, Aβ42, Aβ40, Aβ42/Aβ40 ratio, and Tau/Aβ42 ratio between the AD and control groups (P < 0.05, Table 2). Concentrations of P-tau181 and Tau/Aβ42 ratio were significantly elevated in the AD group compared to the control group. As with Cohort I, the control group had higher concentrations of Aβ42 and the Aβ42/Aβ40 ratio. Interestingly, Aβ40 was significantly higher in the control group than in the AD group, which contrasts with Cohort I where the AD group had higher levels of this indicator. Tau did not significantly differ between the two groups in Cohort II (P = 0.72), whereas in Cohort I, it was significantly higher in the AD group (Figure 2). The combined analysis of both datasets suggests that biomarkers such as P-tau217, P-tau181, Aβ42, Aβ42/Aβ40 ratio, and Tau/Aβ42 ratio display significant differences in AD and hold significant potential for clinical application.

**Table 1.** Basic information on indicators for cohort I.

| | P-tau217 | Aβ40 | Aβ42 | Aβ42/40 | T-tau | T-tau/Aβ42 |
| --- | --- | --- | --- | --- | --- | --- |
| CUC | 0.243±<br>0.133 | 194.925±<br>44.609 | 7.961±<br>2.027 | 0.041±<br>0.008 | 3.066±<br>1.185 | 0.417±<br>0.235 |
| AD | 1.500±<br>0.685 | 220.993±<br>62.910 | 6.555±<br>2.452 | 0.031±<br>0.011 | 3.627±<br>1.640 | 0.695±<br>0.661 |

**Table 2.** Basic information on indicators for cohort II.

| | P-tau181 | Aβ40 | Aβ42 | Aβ42/40 | T-tau | T-tau/Aβ42 |
| --- | --- | --- | --- | --- | --- | --- |
| CUC | 2.038±<br>0.764 | 204.284±<br>65.120 | 15.335±<br>3.256 | 0.083±<br>0.034 | 2.154±<br>0.679 | 0.148±<br>0.063 |
| AD | 3.970±<br>1.387 | 184.417±<br>63.251 | 9.593±<br>2.572 | 0.057±<br>0.021 | 2.188±<br>0.739 | 0.248±<br>0.121 |

**Figure 1.**
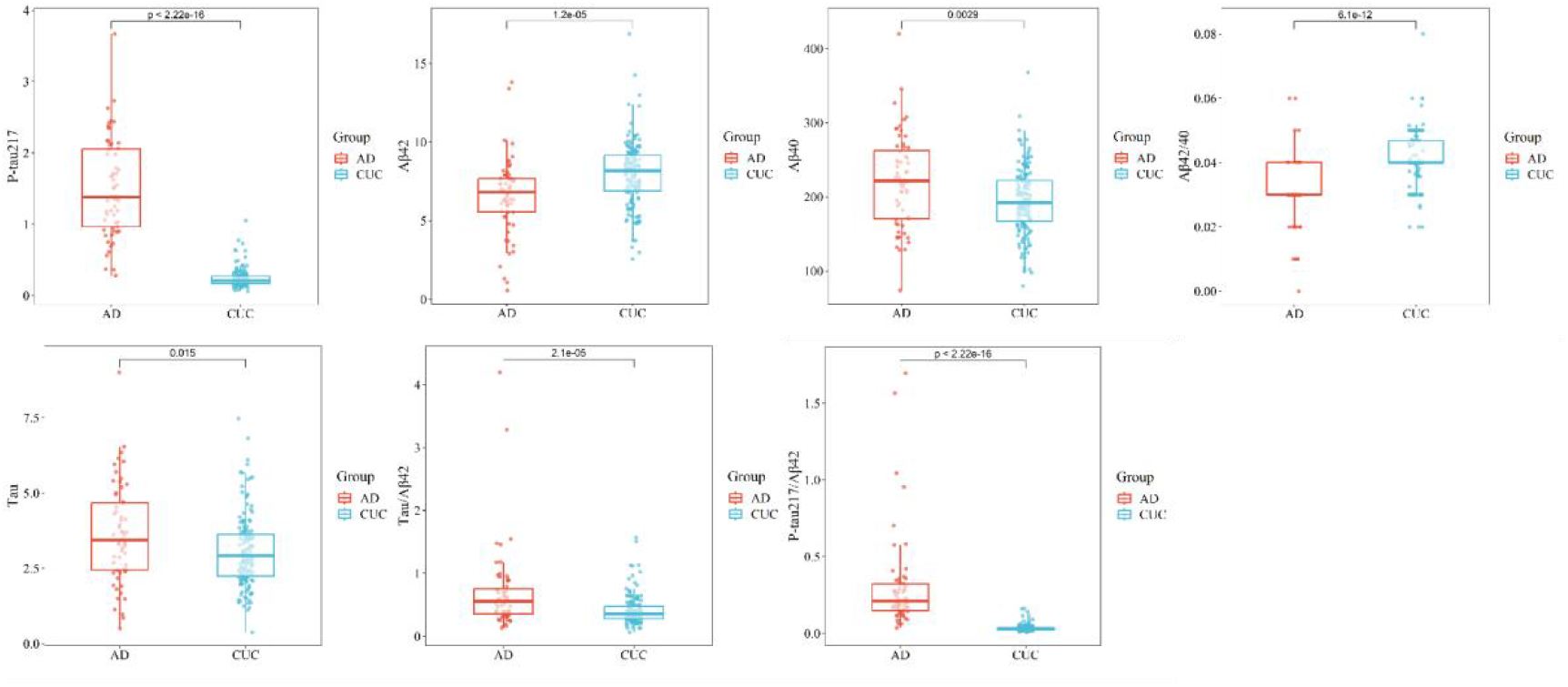
Box plots of concentration distribution of each indicator in different subgroups of Cohort I

**Figure 2.**
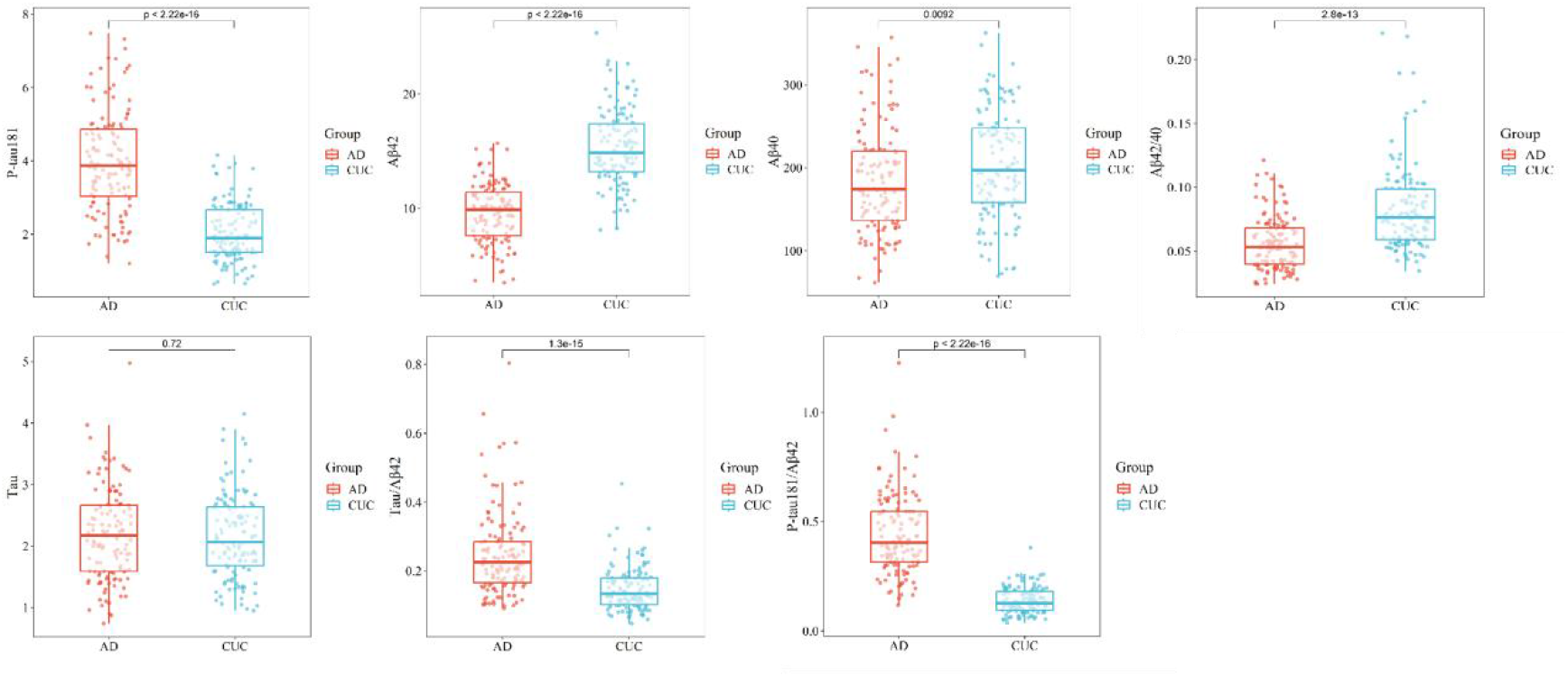
Box plots of concentration distribution of each indicator in different subgroups of Cohort II

### A Comparative Analysis of the Diagnostic Efficacy of Various Indicators in AD

In Cohort I of our study, a detailed comparison was conducted to assess the diagnostic performance of four biomarkers: P-tau217, Aβ42, Aβ42/Aβ40 ratio and Tau/Aβ42 ratio, in both AD and control groups. The resultant data are presented in Table 3. Notably, among these four biomarkers, P-tau217 exhibited a sensitivity of 95.0% and a specificity of 96.0%. Furthermore, its diagnostic accuracy was recorded as 95.7%, marking it as the most precise biomarker among those tested, thereby indicating its ability to effectively differentiate AD from normal controls. In Cohort II, P-tau181 and Aβ42 demonstrated high clinical diagnostic efficacy, with accuracy of 83.1% and 87.1%, respectively (Table 4). However, in Cohort I, Aβ42 displayed a relatively lower accuracy of 62.6%, failing to distinguish the healthy group from the AD group with satisfactory precision. This discrepancy may be attributed to the age difference in the control group between the two cohorts (Cohort I: 50 years old, Cohort II: 70 years old). This variation in age among the control groups across cohorts could have potentially influenced the performance of Aβ42, highlighting the significance of considering age as a confounding factor when evaluating biomarkers. Furthermore, the diagnostic efficacy of each biomarker was evaluated through the computation of the area under the ROC curve (Figure 3). Among all the biomarkers examined, P-tau217 exhibited the highest AUC value (AUC = 0.99), which indicates that this marker holds significant potential in terms of its clinical diagnostic utility. By integrating the data obtained from both Cohort I and Cohort II, it becomes apparent that P-tau217 possesses the highest degree of clinical diagnostic accuracy and emerges as a pivotal biomarker for the clinical diagnosis of AD within the Chinese population.

**Table 3.** Diagnostic performance of each indicator in Cohort I.

| | P-tau217 | Aβ42 | Aβ42/40 | T-tau/Aβ42 |
| --- | --- | --- | --- | --- |
| Accuracy | 95.7% | 62.6% | 82.0% | 70.6% |
| Sensitivity | 95.0% | 81.7% | 71.7% | 60.0% |
| Specificity | 96.0% | 55.0% | 86.1% | 74.8% |
| AUC | 0.990 | 0.693 | 0.793 | 0.688 |

**Table 4.**
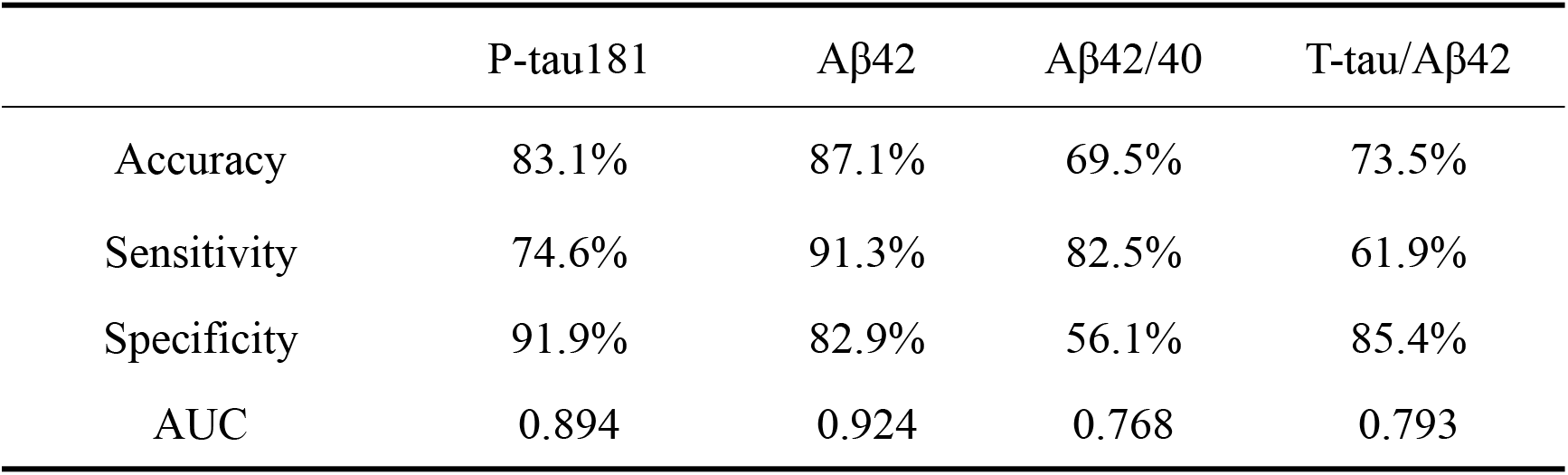
Diagnostic performance of each indicator in Cohort II.

| | P-tau181 | Aβ42 | Aβ42/40 | T-tau/Aβ42 |
| --- | --- | --- | --- | --- |
| Accuracy | 83.1% | 87.1% | 69.5% | 73.5% |
| Sensitivity | 74.6% | 91.3% | 82.5% | 61.9% |
| Specificity | 91.9% | 82.9% | 56.1% | 85.4% |
| AUC | 0.894 | 0.924 | 0.768 | 0.793 |

**Figure 3.**
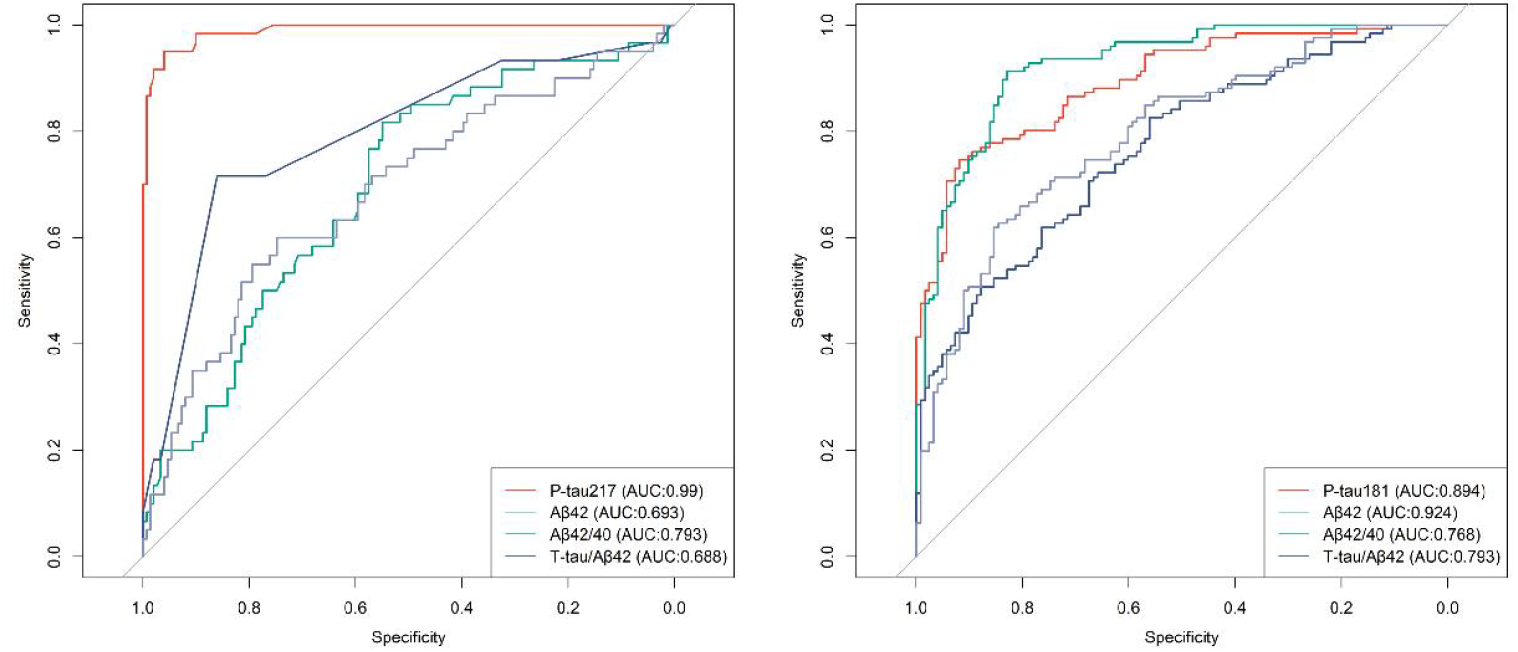
Results of ROC analyses for each indicator in AD vs. controls

## Discussion

In this study, we demonstrated that P-tau181, P-tau217, and Aβ42/Aβ40 ratio all exhibited exceptional performance on the Simoa platform, with statistically significant differences compared to the control group. From an accuracy perspective, P-tau217 was even more advantageous, achieving a concordance with Aβ-PET of up to 95.7% in a multicentre sample from China. It demonstrated outstanding diagnostic performance, and its diagnostic accuracy may surpass that of some approved CSF biomarkers, such as the Tau/Aβ42 ratio (90%) and the Aβ42/Aβ40 ratio (87%), which is comparable to some foreign cohorts^[20]^. In the present study, while some other indicators were significantly different in the two cohorts, they did not exhibit better diagnostic accuracy than P-tau217^[18]^.

Despite the prevalent perception among cognitive experts and clinicians that CSF testing constitutes the gold standard for diagnosis and serves as an alternative to Aβ-PET, this viewpoint has been consistently controversial, as documented in the literature^[21]^. Such controversy may stem from physicians’ inherent caution and the obstacles impeding the dissemination of knowledge^[22]^. However, recent authoritative publications this year have begun to exhibit a gradual shift in this perspective^[18,23]^. Some experts opposes the incorporation of novel blood-based biological indicators in the diagnostic criteria for AD, citing concerns that they may result in misdiagnosis among asymptomatic individuals. Coupled with the current absence of efficacious interventions for asymptomatic individuals with positive markers, this situation may exacerbate unnecessary anxiety and psychological strain. Consequently, a solitary positive biomarker should not be deemed sufficient for a definitive diagnosis of AD^[21]^. Nevertheless, this contention does not impede the pursuit of identifying a more appropriate biomarker, given that similar manifestations are observable in both CSF and Aβ-PET. These similarities may be attributed to an individual’s reserve capacity, potentially leading to the gradual emergence of clinical manifestations of AD at an advanced age, due to various factors^[18,23]^. Moreover, the significance of this concern diminishes in the midst of the profound aging trend prevalent in the country^[3]^. It is also evident that Aβ-PET is limited to detecting neuropathological changes in the middle and late stages of AD^[18]^, and its limited accessibility hinders early diagnosis and treatment of the disease. Several studies have corroborated the effectiveness of plasma P-tau217 in identifying AD, demonstrating superior diagnostic performance compared to CSF markers. This makes blood-based biomarkers a viable candidate for replacing CSF. Furthermore, in China, individuals who test positive for blood markers but exhibit no overt symptoms are classified as high-risk, according to the Chinese consensus on cognitive management. This group should undergo interventions ranging from lifestyle modifications to risk factor management^[24]^, which can substantially mitigate the risk of progressing to AD^[5]^. This approach aligns with the notion of continuous disease management through early prevention and timely treatment.

As the aging population continues to expand, the inadequacy of cognitive centers or memory clinics becomes increasingly apparent when compared to the potentially vast population base in China. According to previous research data, approximately one-quarter of cases may be misdiagnosed when relying solely on symptoms and scores for analysis^[10, 11, 23]^. In the United Kingdom, only approximately 65% of patients with dementia receive a definitive diagnosis, and merely 2% undergo CSF measurements or brain scans for molecular diagnosis. This is due to the frequent lack of capacity in memory clinics to perform such tests^[23]^. Despite the difficulties associated with the development of blood biomarkers, which are present in significantly lower concentrations in the blood compared to CSF, advancements in highly sensitive detection techniques, such as antibodies and mass spectrometry, have enabled the creation of accurate and reliable blood tests. For instance, in a Swedish study involving over 1,200 patients with cognitive impairment, clinicians who incorporated blood test results significantly enhanced diagnostic accuracy to over 90%. This finding suggests that blood biomarkers hold significant potential for applications in the diagnosis of AD^[25]^. It is important to note that these biomarkers do not constitute direct treatments, however, they possess the capability to detect signs of the disease prior to the manifestation of symptoms and brain damage, thereby paving the way for revolutionary therapeutic prospects to delay or even prevent the progression of AD.

Cost-effective blood biomarkers play a key role in AD clinical trials by simplifying participant recruitment, accelerating the study process, and ensuring the accuracy of diagnostic results^[18]^. In addition, these biomarkers help clinicians accurately screen beneficiary patients, effectively monitor treatment effects, and adjust or initiate new treatment regimens when appropriate. The dissemination of this approach will have a profound impact on the early diagnosis and individualised treatment of AD, providing patients with a better prognosis and quality of life.

In the field of AD research, several hematological biomarkers have demonstrated clear correlations, encompassing blood levels of Aβ42, Aβ40, and the Aβ42/Aβ40 ratio^[13]^, phosphorylated tau proteins (P-tau181, P-tau217)^[18, 15, 25]^, as well as plasma concentrations of glial fibrillary acidic protein (GFAP)^[26]^ and neurofilament light chain (NFL)^[27]^. These biomarkers exhibit strong correlations with both cerebrospinal fluid biomarkers and PET results. Specifically, Aβ42, Aβ42/Aβ40, P-tau181, and P-tau217 are recognized as the core biological markers for AD diagnosis, effectively distinguishing the normal population from AD patients. Conversely, due to their lack of specificity, GFAP and NFL are not considered effective differential diagnostic biomarkers^[18, 28]^. Tau proteins play a crucial role in stabilizing neurons within the cell and assisting in the formation of the microtubule skeleton. As AD progresses, tau proteins become more soluble and detach from microtubules, forming more viscous aggregates that exert dual toxicity on neurons. Notably, alterations in tau protein phosphorylation enhance their solubility, and these site-specific phosphorylated tau variants have emerged as the most valuable biomarkers^[23]^. For instance, in the Swedish BioFINDER-2 cohort, an analysis of over 1,400 stored plasma samples revealed that P-tau217 had nearly 100% accuracy in predicting the presence of AD pathology in participants’ brains. This finding was subsequently validated in an independent cohort study published in July 2020^[29]^. These results, in conjunction with our data, suggest that a single biological marker may suffice for the diagnosis of AD, potentially revolutionizing the traditional approach to clinical diagnosis, which relies on cognitive symptoms and the ATN framework. This paradigm shift offers novel perspectives for the early diagnosis of AD and may facilitate earlier interventions and treatment strategies.

There is a prevailing trend in the current scientific community to employ accuracy as the paramount criterion for evaluating the performance of biomarkers. This approach facilitates a more refined categorization of all pertinent biomarkers and aids in making more precise comparisons of their performance. Notably, comparisons of accuracy in Chinese cohorts have yet to be reported. In this academic endeavor, we conducted an investigation on two independent Chinese cohorts, assessing the accuracy of pertinent plasma markers utilizing the Simoa platform for the clinical diagnosis of AD in the Chinese population. Our findings revealed that plasma Aβ42, P-tau181, and P-tau217 exhibited favorable sensitivity and specificity in plasma assays among both AD patients and healthy controls, based on data derived from these two cohorts. When considered as individual indicators, only P-tau217 achieved an accuracy exceeding 90%, thereby qualifying as a standalone indicator for the diagnosis of AD. This observation aligns with the latest international consensus. Furthermore, ratio aspects evaluated in Aβ42/Aβ40 and T-tau/Aβ42 demonstrated varied sensitivities and specificities. These discoveries suggest that these metrics hold potential as diagnostic biomarkers within the Chinese population. Additionally, several other biological markers, such as P-tau181, possess the capability to discern AD patients from controls with high precision and can be utilized as core markers to aid in diagnosis. However, it is acknowledged that the accuracy of these markers, while promising, still requires further enhancement.

For the first time, we conducted an assessment of the accuracy of a single indicator in AD within a Chinese population cohort for evaluative purposes. This approach represents a shift from the previously utilized crude methodology, which assessed indicators based on sensitivity, specificity, or their correlation. Such a shift allows for a valid comparison of the performance of various indicators. The accuracy of these indicators can more effectively highlight their diagnostic value, with a reduced risk of false positives and false negatives. Consequently, they are more likely to meet clinical diagnostic needs, rendering them more suitable for application in primary care centers and large-scale clinical trial screenings. This methodological advancement mitigates the previous overreliance on clinical core indications and composite scales, thereby facilitating earlier identification and diagnosis of AD within the population and enabling systematic management of patients.

Furthermore, our investigation revealed substantial variations in the levels of Aβ42 and the Aβ42/Aβ40 ratio between the two cohorts within the normal population, whereas no such differences were observed in the AD group. A rigorous analysis of our data indicated significant disparities in the age of disease onset among the normal populations included in our study. Specifically, cohort I comprised a younger demographic, whereas cohort II consisted of an older demographic. This age difference may underpin the notable disparities in Aβ42 levels and ratios observed between the two cohorts. Intriguingly, another cohort from China reported similar findings in comparable age groups^[30]^. All three studies employed the Simoa platform assay as their methodological foundation. Comparative analyses of these biomarkers in AD populations with minimal age variation exhibited comparable performance, further corroborating that the observed significant differences are unlikely attributable to variations in reagents or sample preprocessing procedures. By amalgamating the normal populations from the two cohorts, we mitigated the confounding effects of age and regional distribution differences. Additionally, augmenting the sample size will enhance the representativeness of the evaluations conducted on these biological metrics, particularly in the context of their core utility rather than for diagnostic purposes.

Despite our efforts to combine diverse populations to augment the sample size and mitigate the impact of age on the data, it is noteworthy that the diagnostic criteria employed across the samples were not consistent. Specifically, Cohort I utilized Aβ-PET positivity and clinical compliance as the diagnostic criteria, whereas Cohort II relied on cerebrospinal fluid positivity. Additionally, it is plausible that factors beyond clinical asymptomaticity may have played a role in introducing a degree of variability among the patients. In the case of the normal control group, we adopted an imaging-based approach to exclude the possibility of vascular disease or other organic pathologies, thereby minimizing the presence of potential risk factors, such as traumatic brain injury and asymptomatic small-vessel disease, which could elevate the risk of AD. Notably, our analyses within the AD and MCI populations were insufficient to conclusively demonstrate whether there exists a significant difference in the accuracy of these metrics during the early stages of AD (MCI stage). It is anticipated that future meta-analyses or prospective studies with larger sample sizes will provide a more comprehensive assessment of the clinical diagnostic performance of these biological indicators.

## Data Availability

All data produced in the present study are available upon reasonable request to the authors

